# Universal two-dimensional labelled probe-mediated melting curve analysis based on multiplex PCR for rapid typing of plasmodium in a single closed tube

**DOI:** 10.1101/2022.04.12.22271963

**Authors:** Haipo Xu, Xiaolong Liu, Jingfeng Liu

## Abstract

Nowadays, malaria is still one of the major public health problems which commonly caused by four plasmodium species, especially in the epidemic of COVID-19 harboring similar symptoms of fever or fatigue, which easily result in misdiagnosis. The disadvantages of previous traditional detection methods, such as time-consuming, costly, complicated operation, strong professionalism, indistinguishable typing and so on, lead to the dilemma of difficulty to meet the clinical requirements of rapid, easy and accurate typing of common plasmodiums. Herein, we developed and maximally optimized a universal two-dimensional labelled probe-mediated melting curve analysis (UP-MCA) assay based on multiplex PCR for rapid and accurate typing of five plasmodiums, including novel human plasmodium, *Plasmodium knowlesi* (Pk), in a single closed tube following genome extraction. The assay showed the limit of detection (LOD) of 10 copies per reaction and can accurately distinguish plasmodium species from intra-plasmodium and other pathogens. In addition, we also proposed and verified different methods of fluorescence-quenching and two dimensional labelled tag for probes that are suitable for UP-MCA assay. Furthermore, its clinical performance was evaluated by 184 samples and showed sensitivity of 100% (164/164) and specificity of 100% (20/20) at 99% confidence interval, respectively, with the microscopy method as gold standard. Taken together, the UP-MCA system showed excellent sensitivity, specificity and accuracy for genotyping of plasmodium, and it meets the requirements of rapidity and convenience for plasmodium detection in clinical routine and has great potential for clinical translation.

## Introduction

Malaria is a life-threatening disease caused by protozoan parasites, belong to the *Plasmodium* genus, which are transmitted to people through the bites of infected female anopheles mosquitoes. Approximately 229 million malaria cases were reported worldwide according to World Malaria Report 2020, causing an estimated 409,000 deaths in 2019. Although claim of local malaria elimination from some regions or countries authenticated by World Health Organization (WHO), there is still a great risk of imported malaria with the development of economy globalization and the aggravation of population immigration and emigration across regions or countries.

Commonly, there are four parasite species of plasmodium genus known to cause human malaria, including *Plasmodium vivax* (Pv), *Plasmodium falciparum* (Pf), *Plasmodium ovale* (Po), *and Plasmodium malariae* (Pm). Additionally, *Plasmodium knowlesi* (Pk) derived from macaques was considered as the fifth species of Plasmodium causing malaria in humans(1) since large focus and description in 2004(2). Clinically, different treatments and monitoring patterns would be taken after the judgment of different plasmodium species(3). Therefore, timely, easy and accurate typing of plasmodium is very important for countries or coastal cities of happening transactions or population mobility frequently over the world, especially imported malaria from plasmodium high-burden regions.

Traditionally, microscopy, known as the gold standard for the diagnosis of clinical malaria, is professional interpretative and time-consuming, requires expertise gained by strict training and experience, and cannot effectively support large studies. Also, it is difficult in distinguishing *P. knowlesi* from *P. malariae* because of their morphological similarities(2, 4), while *P. knowlesi* results in severe and deadly malaria. Rapid diagnostic tests (RDTs) based on immune-chromatographic antigen detection have been implemented in some diagnostic laboratories as a supplement to microscopy(5). Although they are rapid, simple, and easy to interpret, RDTs target proteins specific to *P. falciparum* or *P. vivax* or those common to all plasmodium species and cannot specifically differ *P. malariae, P. ovale*, and *P. knowlesi*(5, 6). Furthermore, diagnostic sensitivity (e.g., lacking sensitivity for some strains of *P. falciparum* and up to 50% of *P. knowlesi*)(7) and specificity (e.g., *P. vivax* in patients with co-infections and high *P. falciparum* parasitemia levels) are variable widely for commercially available RDTs.

Conversely, molecular detection methods offer an attractive alternative approach. Commonly, PCR is a heat-dependent cycle amplification reaction that harbors ability of exponential amplification for original genome used as one of most important nucleic acid test method(8-10). Conventional PCR assays require post-handling, time-consuming and risk of cross-contamination. Although some real-time PCR assays have been described(11, 12), they cannot distinguish multiple plasmodium species in one-pot reaction mainly as a result of low fluorescent channel and throughput, with similarly limited evaluation of one or two plasmodium species samples. Additionally, clinical performance of specificity cannot be ensured since blood samples from patients with plasmodium are difficult to be obtained and infrequently tested. Recently, Plasmodium species detection assay based on isothermal amplification, such as loop-mediated isothermal amplification (LAMP)(13-16), recombinase polymerase amplification (RPA)(17-19) etc, showed a great convenience for point-of-care diagnosis, especially for resource-limited setting, but it lacks of detection throughput and sensitivity in field diagnosis, especially appearance of false positive or negative results. CRISPR based diagnostic platform (CRISPR-Dx) as a new emerging nucleic acid detection technology used for point-of-care diagnosis, such as Cas12a mediated plasmodium detection assay(20), showed great sensitivity and specificity, but it requires expensive Cas protein from commercial corporation and it is difficult to achieve multiple objects detection in a single closed tube.

Herein, we proposed a Universal two-dimensional labelled Probe-mediated Melting Curve Analysis (UP-MCA) assay based on multiplex PCR within four fluorescence channels at real-time PCR instrument for malaria genotyping. It is a rapid, sensitive, specific, low cost and high throughput detection strategy for five plasmodium species, including *Plasmodium falciparum, Plasmodium vivax, Plasmodium malariae, Plasmodium ovale, and Plasmodium knowlesi*, with *human ribonuclease P* (RNase P) gene as the internal control in state of one-pot and closed tube following introduction of genome extract. Moreover, the assay was validated by detection of clinical malaria samples and showed as a good alternative tool for clinical malaria diagnosis.

## Materials and Method

### Collection and genome extraction for clinical samples

Clinical blood samples were collected from Mengchao Hepatobiliary Hospital of Fujian Medical University from July 2016 to March 2020 with one sample came from one person, including patients with signs and symptoms of, such as fever, fatigue etc. which is similar to malaria. And genome extraction of 200 microliters whole blood for these samples were prepared by Ex-DNA whole blood extraction kit using automated nucleic acid extraction equipment (NP968-C, Xi’an Tianlong Science and Technology Co., Ltd, China) and 80 microliters nucleic acid extract were left over and measured by Nanodrop 2000 (Thermo Fisher Scientific, USA). They were stored at -20 °C until used.

### Principle of species-specific plasmodium detection assay

Although real-time PCR based on sequence specific probe shows higher sensitivity and specificity than that using saturated or unsaturated dye, such as SYBR Green or Eva Green, the amount of detected objects are limited because of limited fluorescent channels of real-time PCR equipment. So, in order to achieve more objects detection in one reaction, we proposed that universal two-dimensional labelled probe-mediated melting curve analysis based on multiplex PCR (UP-MCA) assay for reliable detection of five common species-specific plasmodium in one-pot reaction. This method combines multiplex PCR with melting curve analysis, which is mediated by universal fluorescent probe corresponding to specific fluorescent channel and annealing temperature (Tm) produced for hybridization with homology tag that carried in plasmodium species-specific primer. Its process involves that polymerase mediated asymmetric amplification using target specific primers for tag mark of specific amplicon and fluorescent detection depending on hybridization between homology tag and non-target sequence dependent universal fluorescent probe. Different Tm values brought out by variable degrees of hybridization analyzed by melting curve analysis are used for detection of specific plasmodium candidates in addition to difference of fluorescent channel (Figure.1). According to the previous research, this two dimensional label method can achieve detection of dozens of objects in one reaction under different fluorescence and elaborate Tm value representing detailed detection object(21).

**Figure 1.**
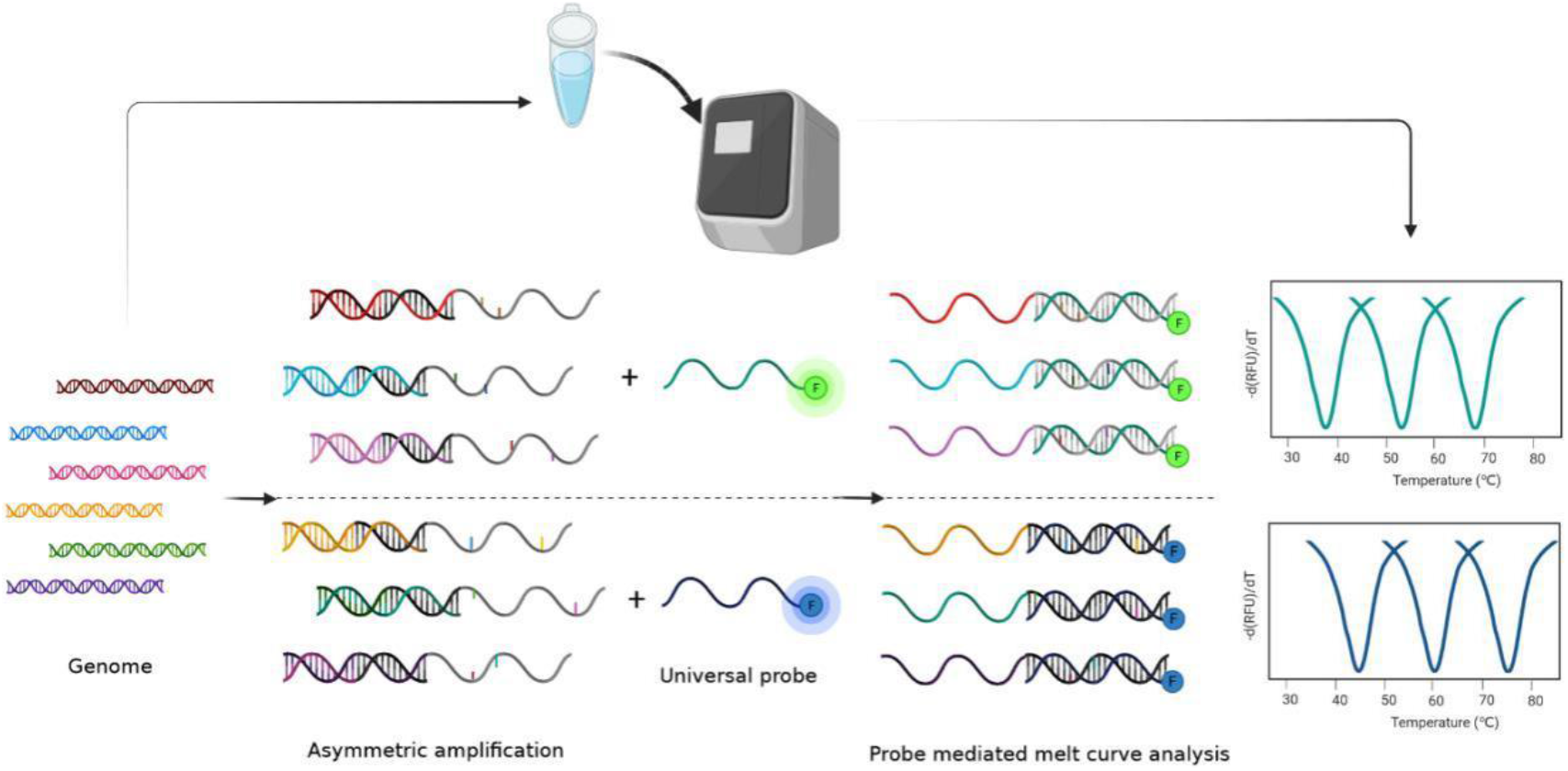
Schematic diagram of universal two-dimensional labelled probe mediated melting curve analysis (UP-MCA) based on multiplex PCR assay for plasmodium species genotyping. Genome of clinical plasmodium sample was put into multiplex PCR reaction for asymmetric amplification and universal fluorescence probe mediated melting curve analysis. The detailed object was judged by fluorescence channel and specific Tm value from melt peak automatically analyzed by instrument software (BioRad CFX Manager software in this paper).

### Design and synthesis of plasmodium species-specific primers and universal probe

Initially, the amplification targets for P. falciparum, P. vivax, P. malariae, P. ovale and P. knowlesi based on previous studies(10, 11, 20, 22-24) and available gene sequences in GenBank were analyzed. Following that, nuclear small subunit (SSU) rRNA gene was confirmed as plasmodium detection target and used for the design of plasmodium species-specific primers as a result of the availability of its sufficient copies and conservative and species-specific sequences for plasmodium detection. Hence, we further blasted the sequences of SSU rRNA gene in P. falciparum (GenBank accession no. M19172), P. vivax (GenBank accession no. X13926), P. ovale (GenBank accession no. L48987), P. malariae (GenBank accession no. M54897), and P. knowlesi (GenBank accession no. AY327550) using the software ClustalX and selected one sequence region existing great difference, also existing high homology for plasmodium genus for the availability of number of primers reduction. Following that, homology and difference sequences were selected for the design of plasmodium UP-MCA assay of abundant forward primer (AF primer) and limiting reverse primer (LR primer), respectively. All primers and fluorescent probes were synthesized from Sunya Biotech (Fuzhou) Co., Ltd and Sangon Biotech (Shanghai) Co., Ltd. And DNA plasmid standards of different plasmodium species-specific and conservative fragments of SSU rRNA gene and fragments of human RNaseP gene were constructed by General Biosystems (Anhui) Co. Ltd.

Universal fluorescent probes and tags for species-specific plasmodium detection were chosen from previous research(21). Furthermore, variable degrees of hybridization between homology tag and universal fluorescent probe determine the detail Tm value resulted from their effective hybridization length and base pairing degree. So we inferred and verified that homologous tags of certain length used for real hybridization with universal probe can also result in different Tm values corresponding to different candidate.

### Establishment of Universal two-dimensional labelled probe-mediated melt curve analysis (UP-MCA) for species-specific plasmodium detection assay

DNA standards of SSU rRNA gene from species-specific plasmodium were prepared before establishment of detection assay (Table. S1). In order to achieve better performance of plasmodium detection, optimizations were implemented by the assay of polymerase enzymes (Taq DNA Polymerase, TaKaRa Taq HS, Vent^®^ (exo-) DNA Polymerase, Klenow Fragment (3’→5’exo-), all of initial concentration are 5 U/μl), buffers (Taq buffer, Taq HS buffer, buffer A, buffer M, buffer 6 and PCR buffer, initial concentrations of all are 10×), temperature (54∼66 °C) and concentration of Mg ^2+^ (3∼6 mM) and primers. Furthermore, Extreme Thermostable Single-Stranded DNA Binding Protein (ET SSB) is a single-stranded DNA binding protein used for stabilization of ssDNA structure and improvement of the processivity of DNA polymerase following that increasing the yield and specificity of PCR. Therefore, we also took ET SSB into consideration for assay optimization.

For single-plex assay, the reaction volume of twenty-five microliter including 1×Taq HS buffer, 1.5 U TaKaRa Taq HS(5 U/μl), 4 mM MgCl_2_, 200 μM dNTP(A/G/C/T), 0.2 μM universal fluorescence probe, 0.8 μM abundant forward primer, 0.04 μM limiting reverse primer with homologous tag, 2 μL template. The assay was performed at the standard two-step PCR protocol with initial denaturation at 95°C for 3 min followed by 50 cycles of denaturation at 95°C for 15 s, annealing / extension at 60 °C fo r 45 s, then addition of melting curve analysis from 35 °C to 85 °C before sufficient denaturation at 95°C for 1 min and hybridization at 30°C for 2 min. Parameter replacement of annealing / extension at 64 °C and addition of 4 ng/uL ET SSB were used for multiplex species-specific plasmodium detection assay. The different channel fluorescence (FAM or HEX or FAM and HEX) can be only collected during the process of melting curve analysis, not for amplification process. The reaction was occurred by the fluorescence PCR equipment (Applied Biosystems 7500 Real-Time PCR System in this study; Thermo Fisher Scientific). As defined herein, the determination of detected result was taken by the fluorescence channel and preset Tm value. And positive result of sample for one subject was refined as the –d(RFU)/dT value of melt peak height was 1.5 times more than that of negative control which RNase-free water or non-plasmodium DNA or RNA serve as reaction template.

### Performance evaluation of UP-MCA assay (including sensitivity, specificity, selectivity and clinical performance) for plasmodium detection

For evaluation of the sensitivity of UP-MCA for species-specific plasmodium assay, the preparation of reaction template was used by a series of gradient dilution concentration of various plasmodium plasmids from 10^5^ copies / μL to 10^1^ copies / μL. Various concentrations of different plasmodium plasmids served as template were run at least in triplex wells. The selection of some pathogens of similar clinical symptoms or blood-borne diseases, including *Babesia, Borrelia burgdorferi, Chikungunya virus*, human immunodeficiency virus, Hepatitis B virus, Hepatitis C virus and *Novel coronavirus (2019)* was applied for the assessment of the specificity of this UP-MCA assay. For examination of the selection and distinction ability of species-specific plasmodium detection assay, two neighboring objects at the same fluorescence channel, *Plasmodium falciparum* and *Plasmodium vivax*, were taken into observation of accurate detection of plasmodium species. Furthermore, negative control of RNase-free water or non-plasmodium genome served as template for plasmodium assay was taken for above three tests.

Practically, the performance evaluation of species-specific plasmodium detection assay was carried out by multiplex UP-MCA fluorescence system using a total of 184 clinical blood samples mentioned above, including 164 microscopy-positive of malaria and 20 microscopy-negative of blood-borne diseases with similar signs and symptoms consistent with malaria. Before they tested, all microscopy results of clinical blood samples were blind for the tester of the assay. And various plasmid standards of objects containing internal control human *RNaseP* gene under the concentration of 100 copies / μL were served as positive control for the assay and the RNase-free water was used as negative control.

### Data analysis

As the analysis of the performance of our UP-MCA assay, we considered microscopy as the gold standard analysis. The discordance of result was considered for samples of which UP-MCA assay detection results did not agree with microscopy results. Discordant samples were retested with alternative multiplex qPCR method based on Taqman probe recommended by World Health Organization(11). Then we considered samples with identical results as true-positive PCR results, since assigning a species is not always possible with microscopy. Moreover, all data were analyzed with GraphPad Prism software. Each experiment was repeated at least three duplicates for each sample.

## Results

### Principle of UP-MCA detection assay for species-specific plasmodium

UP-MCA detection assay is a method that uses asymmetric PCR to achieve the enrichment of two-dimensional labelled products, and combines probe mediated melting curve analysis technology to achieve specific detection of targets with non-target sequence dependent universal fluorescent probes. With regards to asymmetric PCR and melting curve analysis, the difference in the proportion of forward and reverse primers will be introduced into amplification and two-dimensional label should be integrated into amplicon, respectively. Moreover, to further simplify primer design and minify optimization of assay for specificity improvement, we adopted asymmetric PCR of plasmodium genus forward primer which serves as abundant primer, and plasmodium specie-specific reverse primer with homologous tag at the 5’ end representing specific Tm value produced by hybridization with universal probe, which serves as limiting primer for multiplex UP-MCA plasmodium detection (Table 1). These primers were based on target SSU rRNA gene of plasmodium, which its copy number range from 4 to 8 and known to be highly conserved regions suitable for molecular detection of human malaria parasites. This method was also helpful to improve the stability of Tm value of melting curve analysis and avoid the false negative caused by low hybridization between amplicons and specific probes, such as Taqman probe, that are susceptible to mutations of target. For universal fluorescent probe, we proved different kinds of probe quenching method, such as base-quenching and self-quenching, can be used for UP-MCA analysis (Figure.2A). Furthermore, we speculated and also confirmed that different effective lengths of tag could be used for hybridization with probe which was inspired by different Tm value produced by variable degrees of hybridization (via mutations) between homologous tag and fluorescent probe (Figure.2B and Table 1). It seems to be easy to design probes and tags which can simplify a large number of optimization of predicted Tm value via hybrid simulation and synthesis of primer with tag for be tried out. This will help to homogenize temperature range of melting curve analysis and improve detection throughput within a limited temperature range.

**Table 1.**
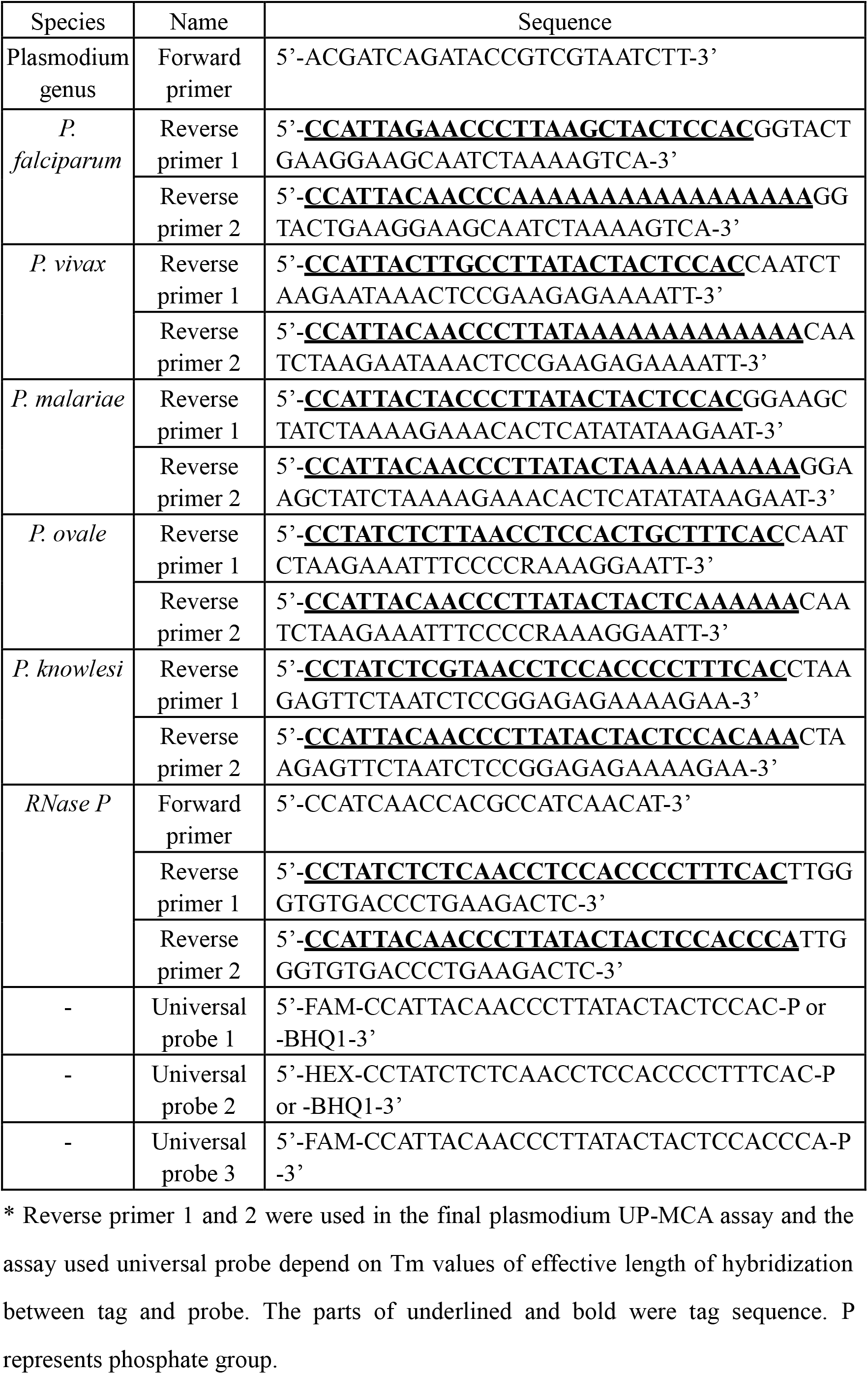
Probes and Primers sequence of UP-MCA assay for plasmodium genotyping.

**Figure 2.**
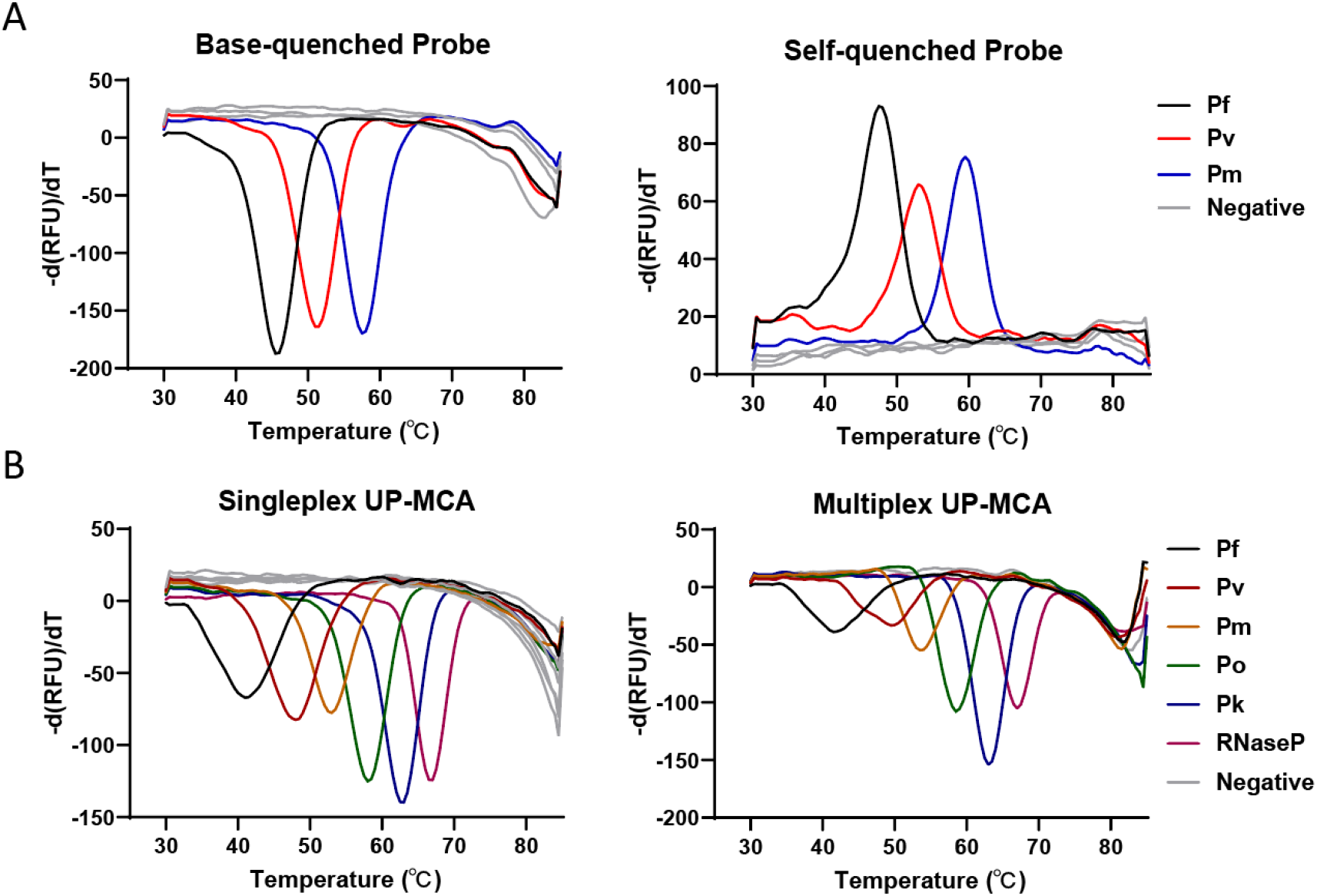
Performance of different kinds of quenched method probe and effective length of hybridization between tag and probe determined Tm value. Performance of base-quenched probe and self-quenched probe for UP-MCA assay. (A). Only one universal probe (universal probe 3) depends on effective length of hybridization between tag and probe for six object UP-MCA detection at a single fluorescence (B).

### Optimization of UP-MCA assay for species-specific plasmodium detection

To improve reaction efficiency, the possible influencing factors were optimized via different enzymes, buffers, reaction temperature and concentration of Mg^2+^. We found that hot-start enzyme of TaqHS and the corresponding buffer or buffer M were the main influencing factors for species-specific plasmodium UP-MCA assay (Figure.3), and the assay obtained higher signal to noise (also difference of melt peak height between positive and negative control) at the parameter of 4 mM Mg^2+^ and 63 °C annealing temperature (Figure.S1 A and B) using the same concentration of target for multiplex plasmodium detection. Interestingly, it seems that ET SSB served as an enhancer of specificity of amplification was not functioned in the multiplex species-specific plasmodium UP-MCA assay (Figure.S1 C).

**Figure 3.**
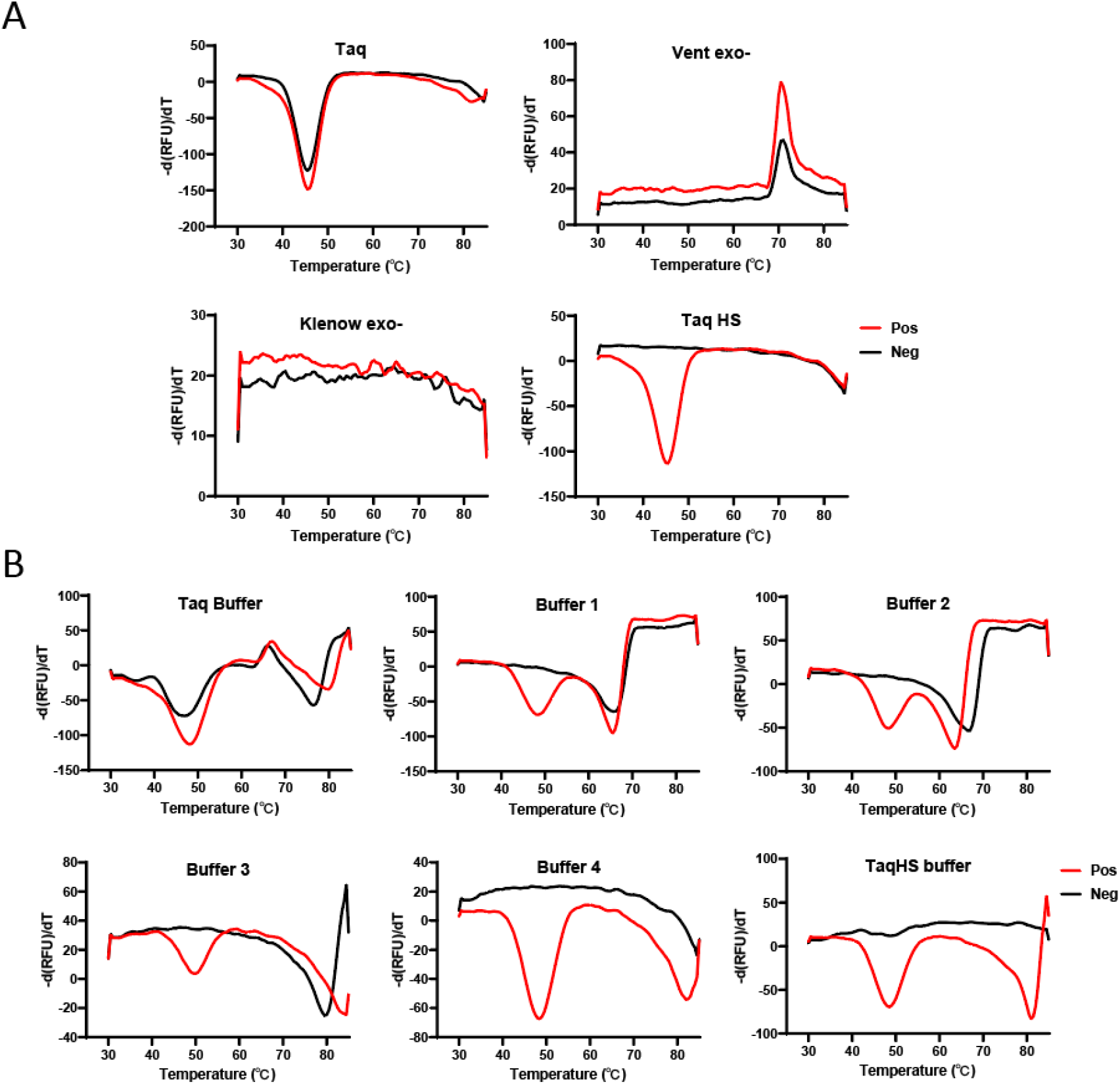
Optimizations of plasmodium UP-MCA assay. Performance of enzymes used for plasmodium UP-MCA assay (A). Performance of different buffers used for plasmodium UP-MCA assay (B).

### Performance of plasmodium UP-MCA detection assay (including sensitivity, specificity, selectivity)

To evaluate the sensitivity of the UP-MCA plasmodium detection assay, a series of gradient concentration of diluted plasmids of plasmodium species and human gene (RNase P, internal control) as reaction template were tested. The results showed that the melt peak height representing negative derivative of fluorescence value with temperature (-d(RFU)/dT) of each object decreases gradually with the decrease of concentration of plasmids, and the LOD for each object was 10 copies / μL (in addition to Pm, 100 copies / μL) (Figure.4A) that nearly access to within the LOD required by WHO.

**Figure 4.**
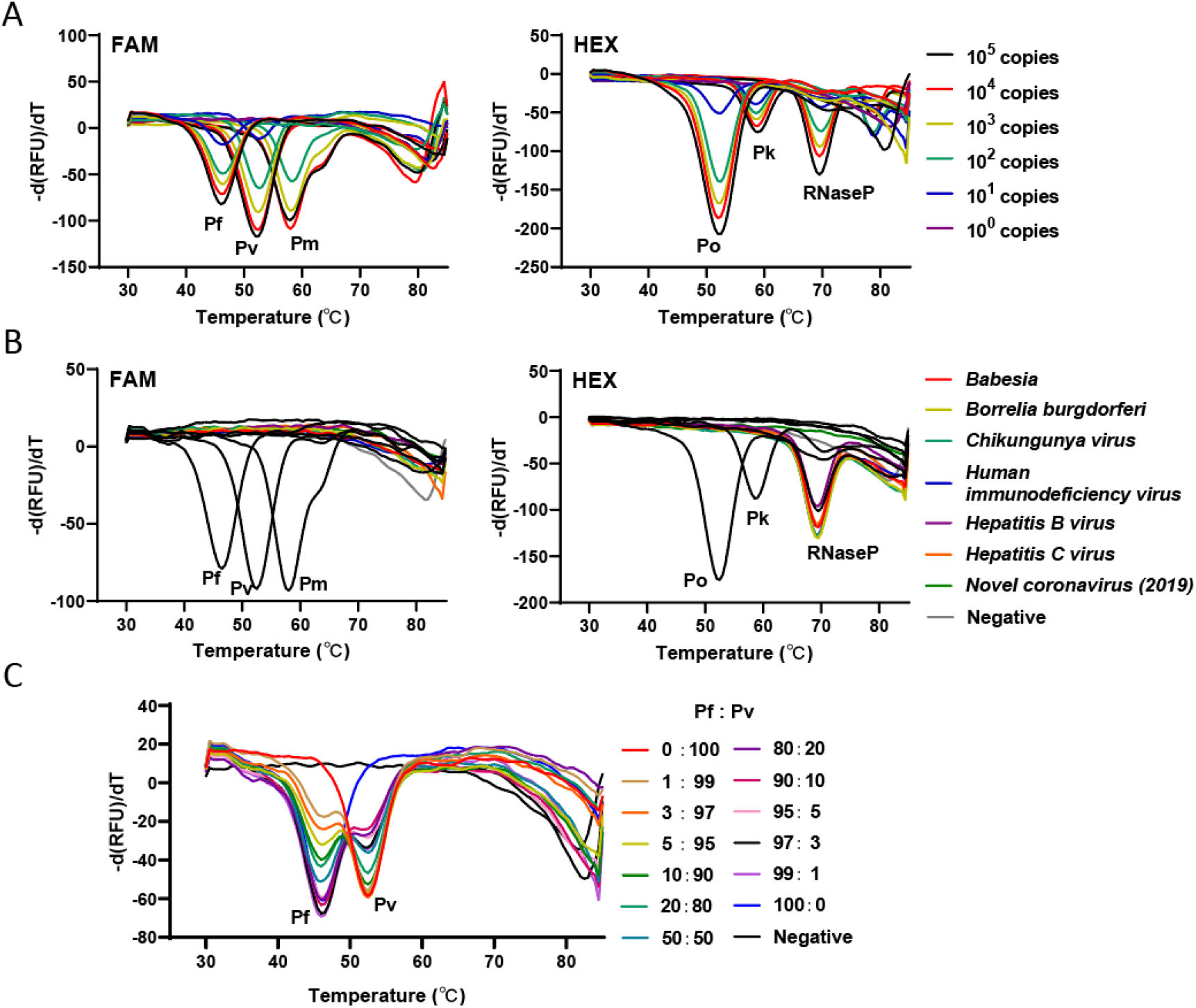
Performance of plasmodium UP-MCA assay. The sensitivity of plasmodium species in the plasmodium UP-MCA assay (A). The specificity of plasmodium UP-MCA assay. The objects used for specificity investigation including Babesia, Borrelia burgdorferi, Chikungunya virus, Human immunodeficiency virus, Hepatitis B virus, Hepatitis C virus, Novel coronavirus (2019) and plasmodium species (B). The selectivity of Plasmodium falciparum and plasmodium vivax in the established plasmodium UP-MCA detection assay (C).

**Figure 5.**
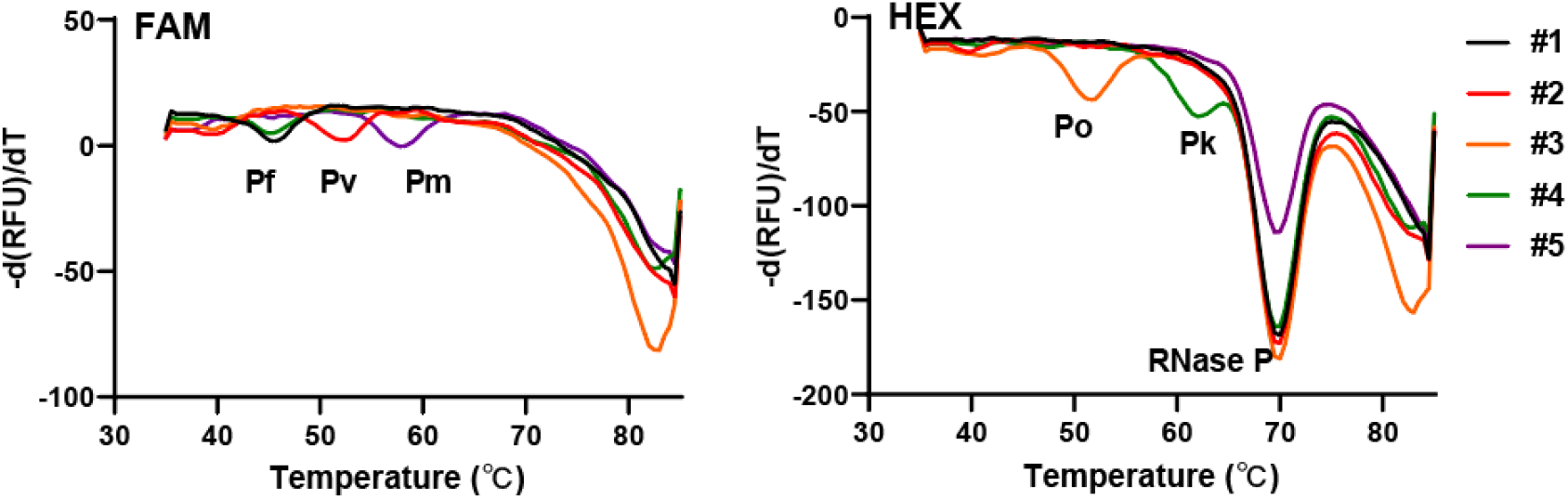
The clinical sample detection example of plasmodium UP-MCA assay.

To assess the specificity of the UP-MCA plasmodium detection assay, genome of samples from a panel of homologous pathogens and that of blood-borne diseases having similar clinical symptoms or signs to malaria were tested. Three duplicates were performed, and 100 copies of species-specific plasmodium standards were used as templates of positive control for each reaction. Consequently, there were no positive results from the tested pathogens except for the DNA standards of species-specific plasmodium (Figure.4B), indicating that the high specificity of species-specific plasmodium UP-MCA assay for detecting plasmodium of detailed genotype.

To investigate the selectivity of the UP-MCA plasmodium detection assay, *Plasmodium falciparum and* its adjacent *Plasmodium vivax* at the same fluorescent channel were selected to test the accuracy of the UP-MCA detection assay. Specifically, different ratios of Pf and Pv plasmid standards under the 100 copies concentration range from 0:100, 1:99, 3:97, 5:95, 10:90, 20:80, 50:50, 80:20, 90:10, 95:5, 97:3, 99:1, 100:0 were prepared and performed by the multiplex plasmodium detection assay. The result showed that the assay exactly detect different kinds of dosage ratios of adjacent detected objects from the same fluorescence channel (Figure.4C). The assay method has excellent discrimination ability of detection.

### Results of clinical sample via UP-MCA detection assay for plasmodium

To evaluate the clinical diagnosis performance of the plasmodium UP-MCA detection assay, 184 patients samples (164 positive-result and 20 negative-result for plasmodium microscopy), which had been confirmed by microscopy served as the gold standard method for malaria, were tested by detector covered microscopical results above. There were 164 plasmodium-positive samples and 20 plasmodium-negative samples detecting by the plasmodium UP-MCA assay which were all consistent to the results of plasmodium microscopy. Compared with the gold standard detection method, the plasmodium UP-MCA assay showed the sensitivity and specificity of both 100% (Table 2).

**Table 2.** Results of UP-MCA assay & microscopy for plasmodium detection.

| Reference method | Results | UP-MCA assay |  | Total |
| --- | --- | --- | --- | --- |
|  |  | Positive | Negative |  |
| Microscopy | Positive | 164 | 0 | 164 |
|  | Negative | 0 | 20 | 20 |
| Total |  | 164 | 20 | 184 |

## Discussion

PCR, as one of the most widely used molecular biology detection methods, has the great advantages of high sensitivity and specificity, simplicity and rapidity, and low cost, especially for real-time quantitative PCR. It has been used in many fields including pathogen detection, tumor marker detection, and genetic identification and so on. We demonstrated universal two-dimensional labelled probe-mediated melting curve analysis (UP-MCA) assay based on multiplex PCR for malaria that are sensitive and specific for detection of the 5 major plasmodium species as well as the human *RNase P* gene as an internal reference control with the employment of two fluorescence channel. The multiplex plasmodium PCR assay can be performed rapidly (nearly 3 hours, including 1 hour for genome extraction and 2 hours for amplification and detection) in a single closed tube, and the 384-well or 96-well format both achieve high detection throughput, which is required for large clinical sample detection and epidemiologic studies of imported malaria.

We rigorously investigated the plasmodium target gene based on its inherent characteristics (including conservation, specificity and copy number etc.) and previous published studies. The nuclear small subunit (SSU) rRNA gene of plasmodium harboring four to eight copy numbers was carefully defined as target gene for the reason that not only that is known to be highly conserved regions suitable for plasmodium-genus primers selection, but also its existence of plasmodium-species region for specific primers design. Less optimization can be conducted by one abundant primer selected from conservative region and numbers of limiting primers designed from species-specific region for multiplex and asymmetric PCR detection for plasmodium. Furthermore, many optimizations for detection efficiency improvement were rigorously carried out, including main factors of specific primers, enzymes and best matching buffer etc.

For the UP-MCA assay, engineered fluorescent probes were not directly specific for target sequence, but it was dependent on species-specific limiting primer used in asymmetric PCR resulting in abundant single-strand DNA products with specific tag for special hybridization. It is helpful to improve the resistance of mutations or SNP existed in probe hybridization region and theoretically increase sensitivity of the plasmodium detection assay. Moreover, in addition to mutation existing in homologous tag, our experiment showed different effect hybridization length between tag and fluorescent probe also can be applied in two-dimensional label design strategies for universal probe. It was great advantage for difficulty reduction of two-dimensional label design and throughput improvement of detection target following homogenization of melting temperature, unfortunately lacking of further more detection objects in one-pot to be validated.

Limitations were also demonstrated in the plasmodium UP-MCA detection assay. Firstly, there should be more targets to be validated for detection throughput improvement used in plasmodium UP-MCA assay, and less detection objects and numbers of primers may be the reason of ET SSB seldom seems to be functioned. But, in other words, primers designed and used in the assay were adequately specific for species-specific plasmodium. Secondly, the UP-MCA assay was one of half-quantitative multiplex method that could not accurately achieve to quantitative plasmodium species. Furthermore, more clinical samples collected from multicenter should be incorporated into the assay for clinical performance validation.

In summary, we developed one plasmodium species-specific detection assay for greatly enhancing sensitivity and specificity that was important for import malaria derived from regions or cities of frequent communications for economy or population emigration and immigration. It can be rapidly and accurately genotype 5 plasmodium species and great helpful to increase the ability of eliminating imported malaria. In summary, the developed plasmodium UP-MCA assay exhibited excellent sensitivity, specificity and selectivity for plasmodium species-specific detection. It is helpful to rapid and sensitive genotyping of 5 species-specific plasmodiums in a single closed tube under malaria control plan accordingly proposed by World Health Organization.

## Data Availability

All data produced in the present work are contained in the manuscript.

## Supplemental file

**Table.S1.**
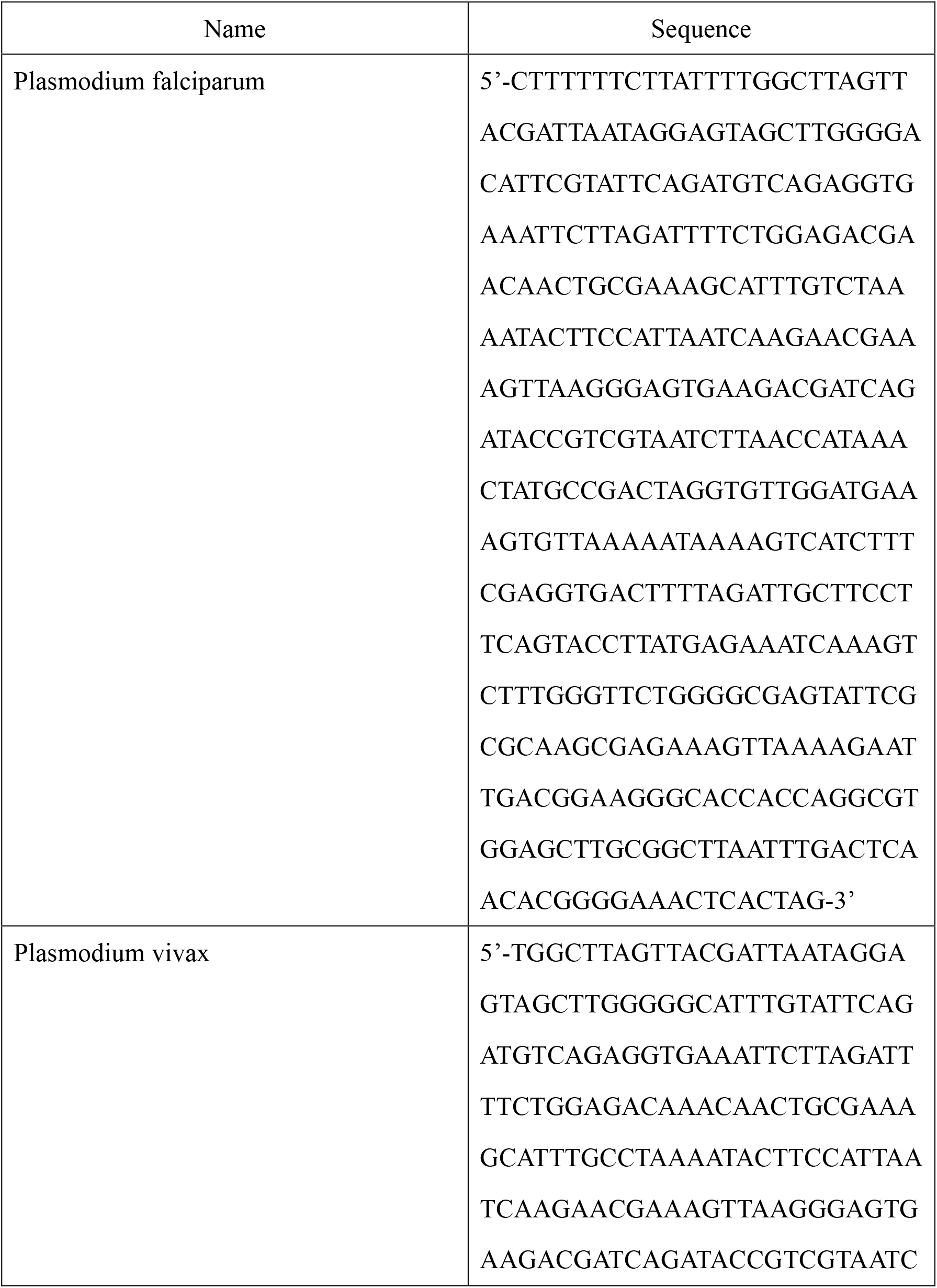
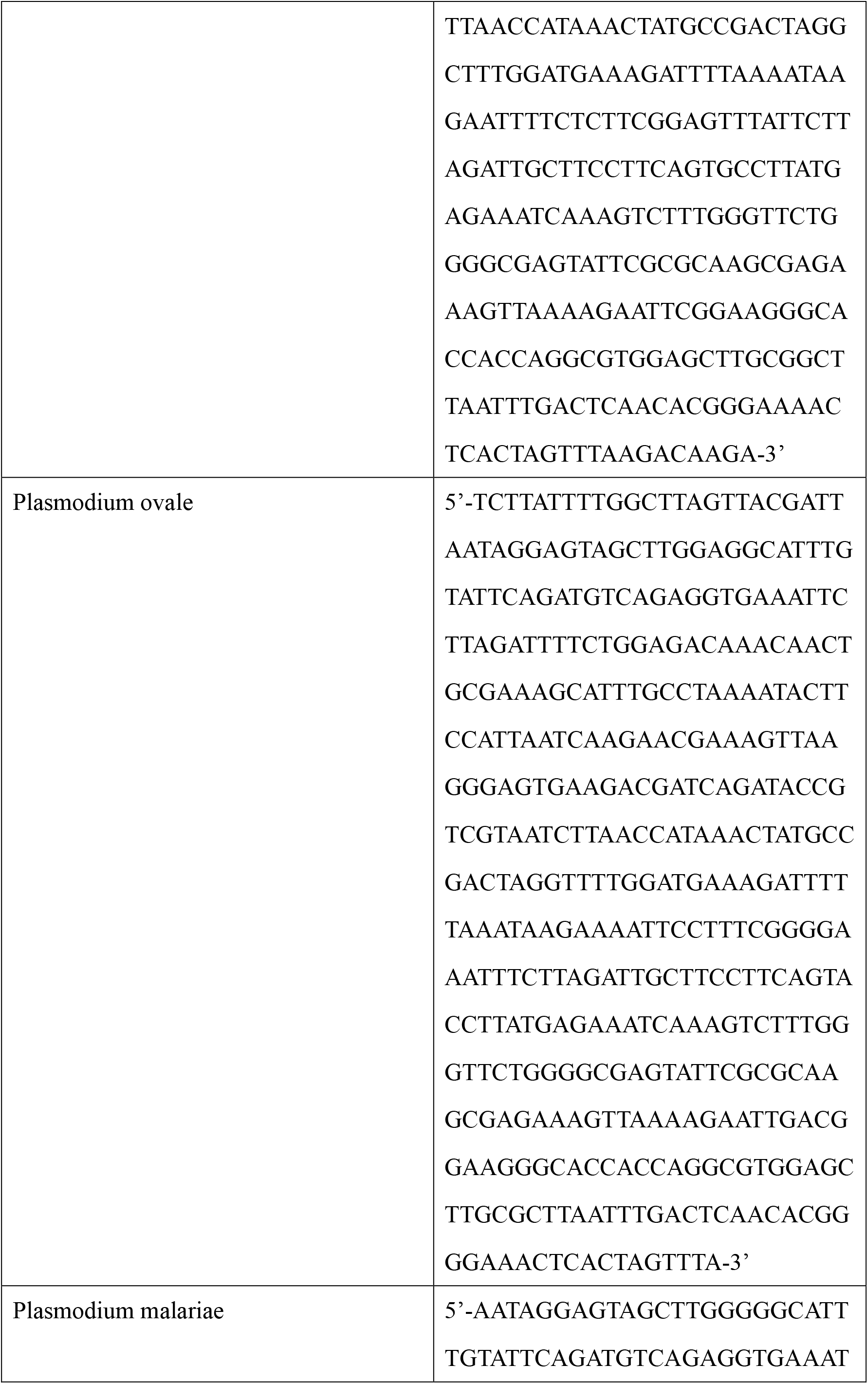
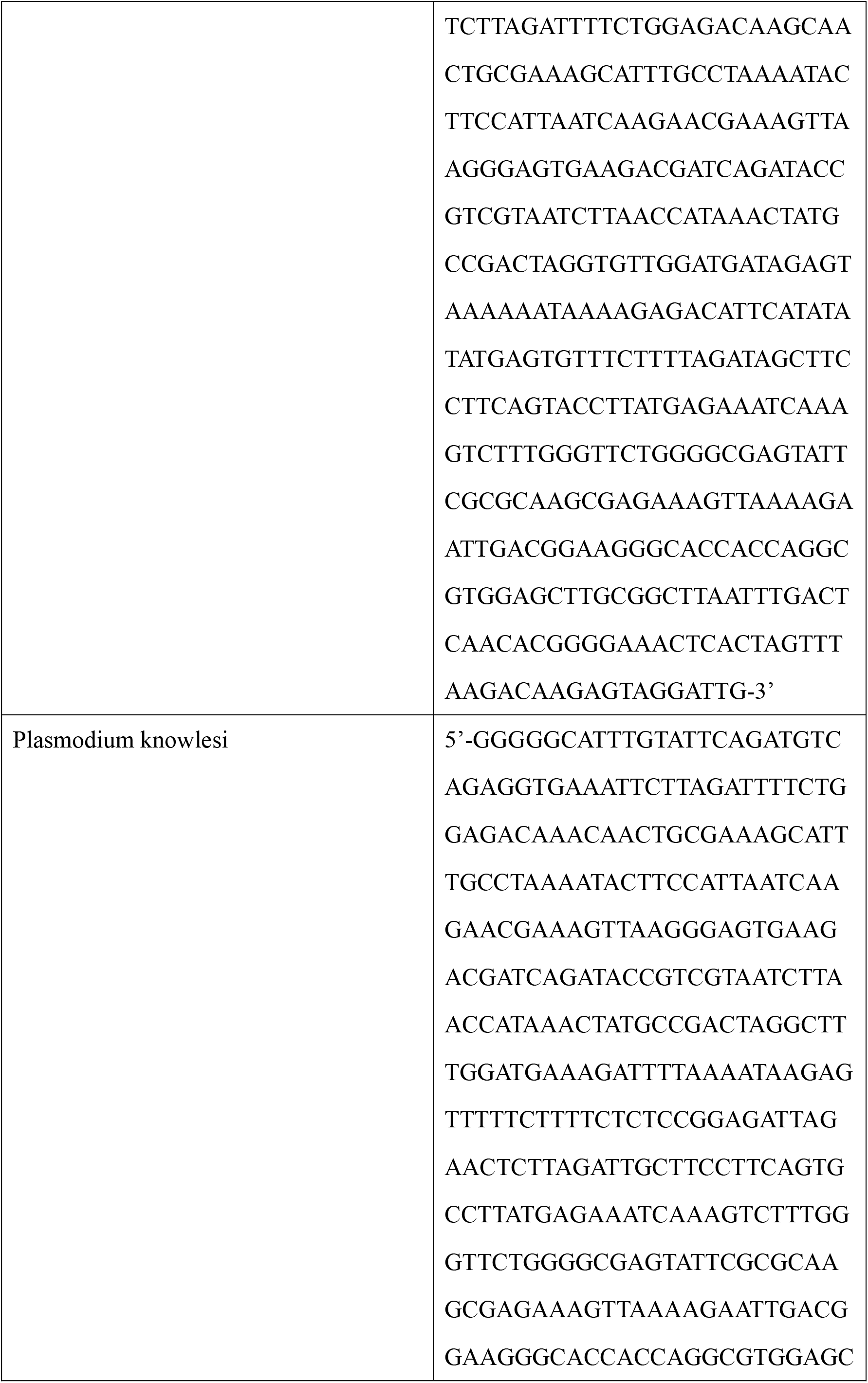
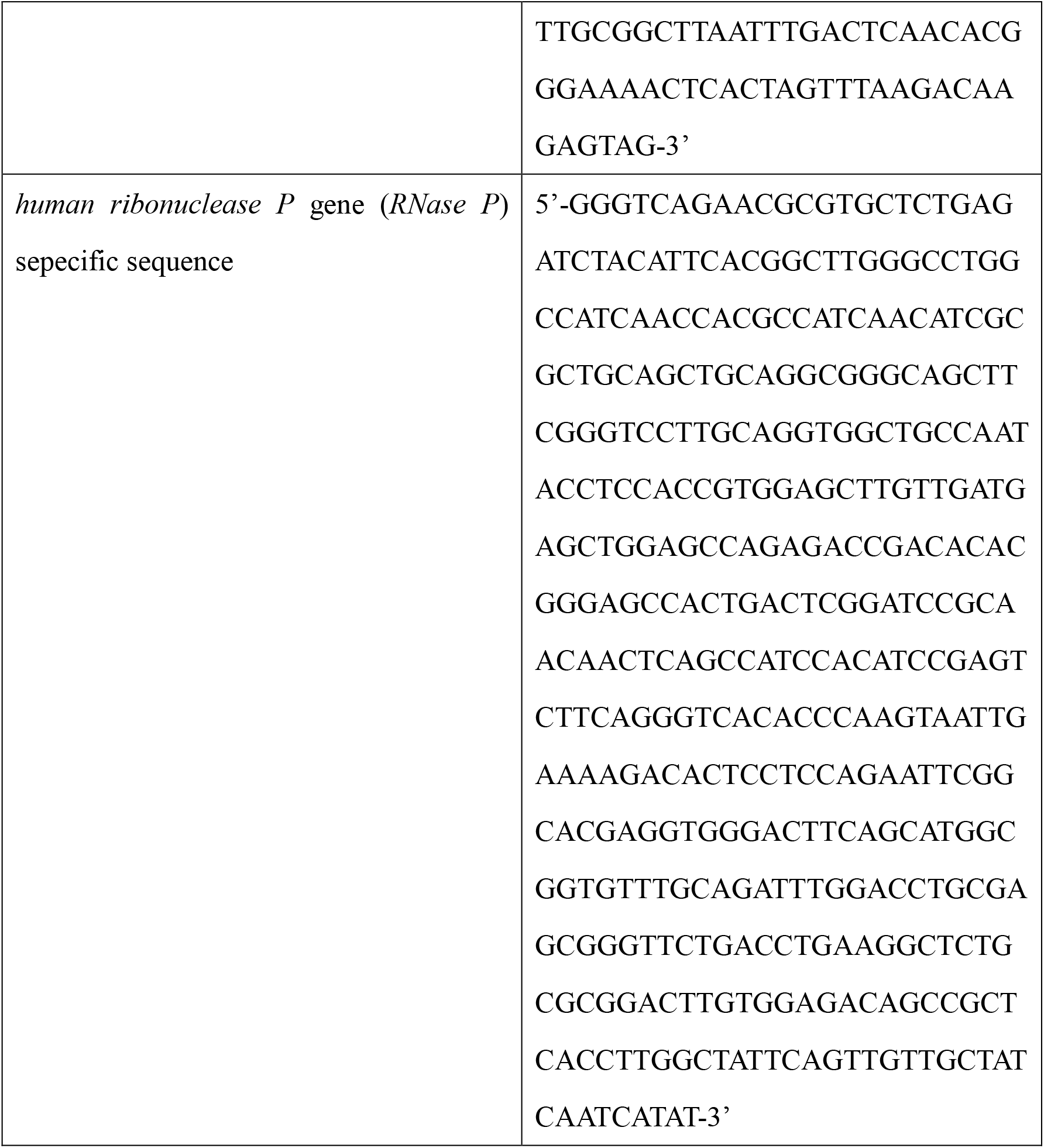
Nucleic acid sequence of SSU rRNA gene from species-specific plasmodium or *human ribonuclease P* gene embedded in PUC57 vector for plasmid standards.

## Figure Legends

**Figure.S1.**
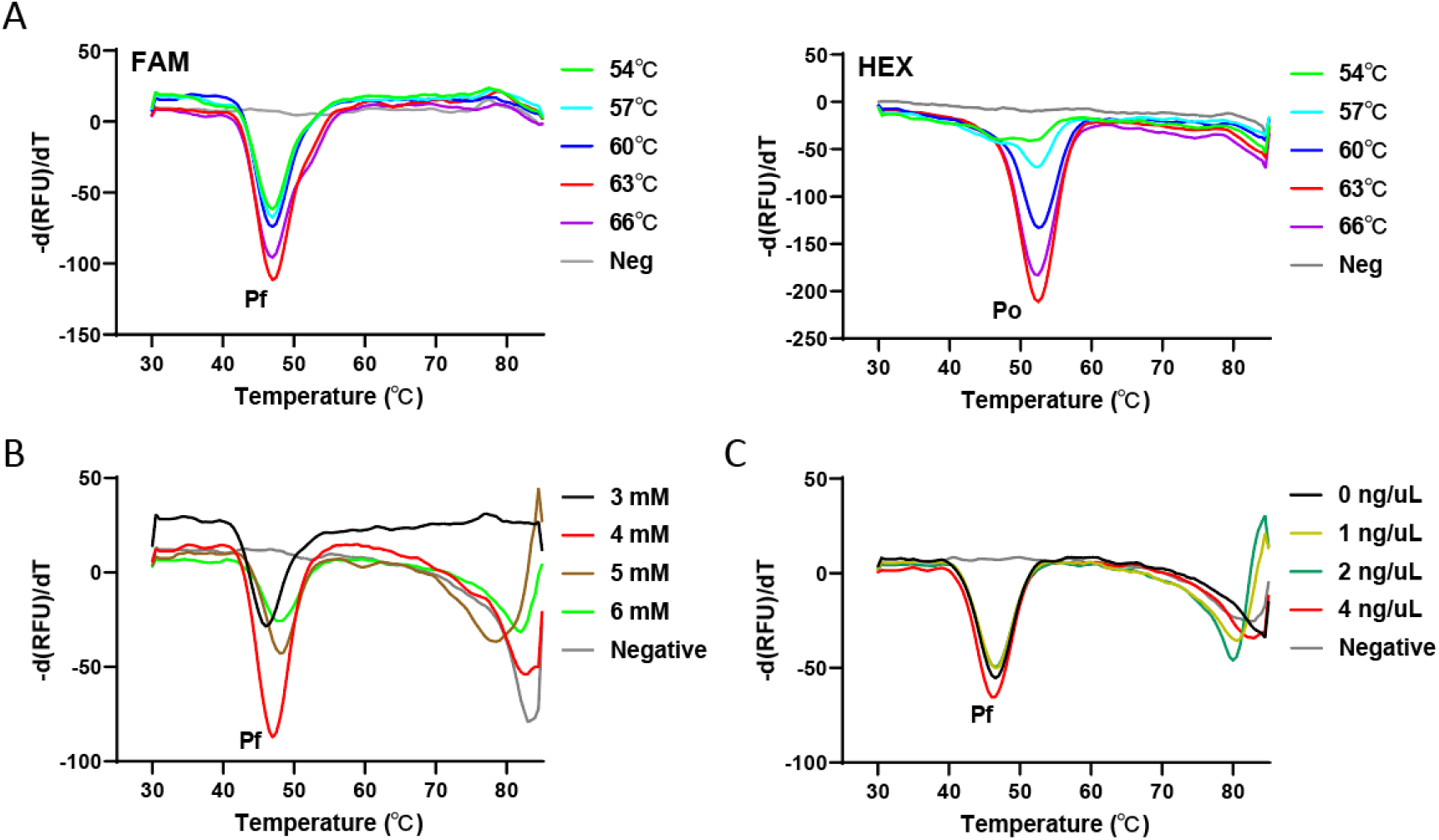
Optimizations of plasmodium UP-MCA assay. Detection performance of plasmodium UP-MCA assay at different annealing temperature (A). The efficiency of plasmodium UP-MCA assay at different concentration of Mg ion (3 mM, 4 mM, 5 mM, 6 mM) (B). The amplification efficiency for plasmodium UP-MCA detection assay at different concentration of ET SSB (C).

## Notes

**Funding:** Supported by Natural Science Foundation of Fujian Province (Grant No. 2021J011279), Scientific Foundation of Fujian Health Department (Grant No. 2019-1-87), Scientific Foundation of Fuzhou Science and Technology Department (Grant No. 2019-S-90), Startup Fund for scientific research, Fujian Medical University (Grant No. 2019QH1299).

### Competing Interest Statement

The authors have declared no competing interest.

### Funding Statement

This study was funded by Fujian Provincial Clinical Research Center for Hepatobiliary and Pancreatic Tumors (Grant No. 2020Y2013), Natural Science Foundation of Fujian Province (Grant No. 2021J011279), Scientific Foundation of Fujian Health Department (Grant No. 2019-1-87), Startup Fund for scientific research, Fujian Medical University (Grant No. 2019QH1299).

### Author Declarations

Ethics committee of Mengchao Hepatobiliary Hospital of Fujian Medical University gave ethical approval for this work.

## Reference

1. White NJ. 2008. Plasmodium knowlesi: the fifth human malaria parasite. Clin Infect Dis 46:172–3.

2. Singh B, Kim Sung L, Matusop A, Radhakrishnan A, Shamsul SS, Cox-Singh J, Thomas A, Conway DJ. 2004. A large focus of naturally acquired Plasmodium knowlesi infections in human beings. Lancet 363:1017–24.

3. Organization WHJW. 2015. Guidelines for the Treatment of Malaria. Third Edition (PDF).

4. Singh B, Daneshvar C. 2013. Human infections and detection of Plasmodium knowlesi. Clin Microbiol Rev 26:165–84.

5. Wilson ML. 2012. Malaria rapid diagnostic tests. Clin Infect Dis 54:1637–41.

6. Erdman LK, Kain KC. 2008. Molecular diagnostic and surveillance tools for global malaria control. Travel Med Infect Dis 6:82–99.

7. Barber BE, William T, Grigg MJ, Piera K, Yeo TW, Anstey NM. 2013. Evaluation of the sensitivity of a pLDH-based and an aldolase-based rapid diagnostic test for diagnosis of uncomplicated and severe malaria caused by PCR-confirmed Plasmodium knowlesi, Plasmodium falciparum, and Plasmodium vivax. J Clin Microbiol 51:1118–23.

8. Putaporntip C, Buppan P, Jongwutiwes S. 2011. Improved performance with saliva and urine as alternative DNA sources for malaria diagnosis by mitochondrial DNA-based PCR assays. Clin Microbiol Infect 17:1484–91.

9. Canier L, Khim N, Kim S, Sluydts V, Heng S, Dourng D, Eam R, Chy S, Khean C, Loch K, Ken M, Lim H, Siv S, Tho S, Masse-Navette P, Gryseels C, Uk S, Van Roey K, Grietens KP, Sokny M, Thavrin B, Chuor CM, Deubel V, Durnez L, Coosemans M, Ménard D. 2013. An innovative tool for moving malaria PCR detection of parasite reservoir into the field. Malar J 12:405.

10. Komaki-Yasuda K, Vincent JP, Nakatsu M, Kato Y, Ohmagari N, Kano S. 2018. A novel PCR-based system for the detection of four species of human malaria parasites and Plasmodium knowlesi. PLoS One 13:e0191886.

11. Rougemont M, Van Saanen M, Sahli R, Hinrikson HP, Bille J, Jaton K. 2004. Detection of four Plasmodium species in blood from humans by 18S rRNA gene subunit-based and species-specific real-time PCR assays. J Clin Microbiol 42:5636–43.

12. Dos Santos EH, Yamamoto L, Domingues W, di Santi SM, Kanunfre KA, Okay TS. 2020. A new Real Time PCR with species-specific primers from Plasmodium malariae/P. brasilianum mitochondrial cytochrome b gene. Parasitol Int 76:102069.

13. Piera KA, Aziz A, William T, Bell D, González IJ, Barber BE, Anstey NM, Grigg MJ. 2017. Detection of Plasmodium knowlesi, Plasmodium falciparum and Plasmodium vivax using loop-mediated isothermal amplification (LAMP) in a co-endemic area in Malaysia. Malar J 16:29.

14. Hashimoto M, Sakamoto H, Ido Y, Tanaka M, Yatsushiro S, Kajimoto K, Kataoka M. 2018. In situ loop-mediated isothermal amplification (LAMP) for identification of Plasmodium species in wide-range thin blood smears. Malar J 17:235.

15. Nolasco O, Montoya J, Rosales Rosas AL, Barrientos S, Rosanas-Urgell A, Gamboa D. 2021. Multicopy targets for Plasmodium vivax and Plasmodium falciparum detection by colorimetric LAMP. Malar J 20:225.

16. Tambo M, Auala JR, Sturrock HJ, Kleinschmidt I, Bock R, Smith JL, Gosling R, Mumbengegwi DR. 2018. Evaluation of loop-mediated isothermal amplification as a surveillance tool for malaria in reactive case detection moving towards elimination. Malar J 17:255.

17. Lalremruata A, Nguyen TT, McCall MBB, Mombo-Ngoma G, Agnandji ST, Adegnika AA, Lell B, Ramharter M, Hoffman SL, Kremsner PG, Mordmüller B. 2020. Recombinase Polymerase Amplification and Lateral Flow Assay for Ultrasensitive Detection of Low-Density Plasmodium falciparum Infection from Controlled Human Malaria Infection Studies and Naturally Acquired Infections. J Clin Microbiol 58.

18. Lai MY, Lau YL. 2020. Detection of Plasmodium knowlesi using recombinase polymerase amplification (RPA) combined with SYBR Green I. Acta Trop 208:105511.

19. Lai MY, Ooi CH, Lau YL. 2018. Recombinase Polymerase Amplification Combined with a Lateral Flow Strip for the Detection of Plasmodium knowlesi. Am J Trop Med Hyg 98:700–703.

20. Lee RA, Puig H, Nguyen PQ, Angenent-Mari NM, Donghia NM, McGee JP, Dvorin JD, Klapperich CM, Pollock NR, Collins JJ. 2020. Ultrasensitive CRISPR-based diagnostic for field-applicable detection of Plasmodium species in symptomatic and asymptomatic malaria. Proc Natl Acad Sci U S A 117:25722–25731.

21. Zhan Y, Zhang J, Yao S, Luo G. 2020. High-Throughput Two-Dimensional Polymerase Chain Reaction Technology. Anal Chem 92:674–682.

22. Reller ME, Chen WH, Dalton J, Lichay MA, Dumler JS. 2013. Multiplex 5’ nuclease quantitative real-time PCR for clinical diagnosis of malaria and species-level identification and epidemiologic evaluation of malaria-causing parasites, including Plasmodium knowlesi. J Clin Microbiol 51:2931–8.

23. Nuin NA, Tan AF, Lew YL, Piera KA, William T, Rajahram GS, Jelip J, Dony JF, Mohammad R, Cooper DJ, Barber BE, Anstey NM, Chua TH, Grigg MJ. 2020. Comparative evaluation of two commercial real-time PCR kits (QuantiFast™ and abTES™) for the detection of Plasmodium knowlesi and other Plasmodium species in Sabah, Malaysia. Malar J 19:306.

24. Lee PC, Chong ET, Anderios F, Al Lim Y, Chew CH, Chua KH. 2015. Molecular detection of human Plasmodium species in Sabah using PlasmoNex™ multiplex PCR and hydrolysis probes real-time PCR. Malar J 14:28.

